# Religion and COVID 19 Pandemic for better or for worse?

**DOI:** 10.1101/2024.09.04.24312776

**Authors:** Sameera Upashantha Ranasinghe, Lakshitha Iroshan Ranasinghe, Indika Pathiraja

**Affiliations:** Ministry of Health, Sri Lanka; Family Health Bureau,Sri Lanka

## Abstract

COVID-19 has negative repercussions on psychological aspects of the individual. Provision of personalized assistance to each individual is a task which cannot be fulfilled by the health system. In this context religion remains to be an effective tool. A qualitative study was conducted to find the spiritual practices and coping mechanisms of COVID-19 infected people(N=15, mean age =46.3 years) in Kurunegala district. Data collection was conducted using in-depth interviews and thematic analysis was conducted. The main themes found were Fatalism and perceived risk of COVID 19, Adherence to health care guidelines despite fatalistic beliefs, Religious coping in COVID 19, Spiritual practices for coping with stress/ distress and Fatalism as a positive force for self-care. It is imperative to manage the pandemic with the assistance of religion and spiritual practices.

## Introduction

COVID 19 has impacted not only the health aspects but also many socio-economic conditions in societies around the world. On the individual level this pandemic has imposed many challenges. COVID-19 infection is a novel disease with an unusual spread. It has led to a boom of individual perceptions and attitudes among people, which have mostly not been scientifically proven. This uncertainty made people deviate from adhering to health guidelines and their psychological response to the disaster situation. Such individual perceptions are molded through many external and individual influences. Religion is one of the most prominent modifiers of disaster response.

Religion and spirituality are parts of a particular culture. (1, 2). Religiosity is defined as the compliance to religious practices and identifying belonging to a religion. Spirituality is defined as the individual practicing of the religious practices and belief systems(3).

Numerous research depicts that religion and spirituality are associated with physical and psychological wellbeing(4, 5). Especially during disastrous times like pandemics, people are drawn more towards their religion(6). Direct exposure to COVID 19 infected patients was associated with high religiosity and high religiosity with less stress level(7). Religion has been found to be an effective tool to cope with the psychological crisis associated with COVID-19 infection (8, 9). Religious leaders have been used as conveyers of health messages, stress relievers and mediators between health officials and the public during COVID 19 pandemic(10). A hotline providing spiritual and religious guidance to multireligious people who are distressed at COVID 19 pandemic has been established in Brazil(11). It was revealed that the spiritual beliefs aided the public to relieve mental distress during the COVID 19 pandemic (12).

Even though the modern health care systems are capable of substantially controlling epidemics at the country level, the results of which appear significant on paper have failed to successfully address the epidemic’s undesirable effects on the individual. Providing individual level guidance to a population puts a huge burden on a health system that cannot be achieved through a health system alone during a pandemic. Thus the use of religion as a tool to address the above health-related problems could yield successful results.

Asian societies are different from western societies in many ways(13, 14). These differences are marked mainly in *‘religiosity’* and other moral values in-between Asia and the Western world. (15)The Sri Lankan society is complex with a religious landscape with most Buddhists and a minority of Islams, Hindus and various sects of Catholicism. The employment of religion as a viewpoint and a medium of coping with the COVID 19 can have different implications than the outside world.

## Methodology

The objective of the research is to find the spiritual practices and coping mechanisms of COVID 19 infected people in Kurunegala district. The sample consisted of people infected with COVID 19 and being treated for COVID 19 at treatment centers in the Kurunegala District. A total of 15 patients were interviewed, and the mean age of the participants was 46.3 years. All the study participants had been treated at the treatment center for more than one week (for the purpose of involving people who have been with the condition for some time).

In-depth interviews were conducted among the participants in their native language through video conversations. The participants were interviewed while on treatment or while receiving intermediate care. Each interview lasted approximately 20 minutes. Two investigators took notes on the verbal and non-verbal expressions. Data and investigator triangulation methods were used to improve the validity of the data.

The transcriptions were then translated, back-translated, and accounted for any discrepancies. The final agreed English transcription was analyzed using RQDA software to identify codes and thematic analysis henceforth.

Open ended questions

1. How would you describe your spiritual self?/ Do you consider yourself to be a religious person?
2. Were you afraid that you might get infected with COVID 19?
3. How has your spirituality or religiosity helped you to cope during the COVID-19 crisis?
4. How do you believe that past events have impacted your resilience after being infected with COVID-19?-Faith in god or else
5. What has given you hope in the midst of the infection period?-God, kindness or fate

## Results

Socio demographic details of the participants

**Table 1:** describes the socio demographic details of the participants.

| Variable | Category | Count | Percentage |
| --- | --- | --- | --- |
| Age | 30-40 | 5 | 33% |
|  | 41-50 | 4 | 27% |
|  | 51-60 | 6 | 40% |
| Sex | Female | 9 | 60% |
|  | Male | 6 | 40% |
| Educational status | Primary | 2 | 13% |
|  | Secondary | 3 | 20% |
|  | Pass Ordinary Level | 2 | 13% |
|  | Pass Advanced level | 6 | 40% |
|  | Graduate | 1 | 7% |
|  | Post Graduate | 1 | 7% |

Out of the participants majority (40%), belonged to 51-60 age group, female sex (60%) and who have passed Advanced Level (40%).

### Themes

We generated the following themes for this study.

#### 1. Fatalism and perceived risk of COVID 19

The majority of the participants have expressed that they strongly believed that they would never acquire the infection. They had stated that they had trusted higher power or order protects them. This higher power could be in various forms from; God, fate or Karma.

Through their spiritual practice, they had a belief that these powers could protect them from acquiring COVID 19 infection.

*“I always observe ‘Pansil’ and donate to charity. I never even kill a mosquito. I never thought that this could happen to me*.*”*

*“I prayed to God to save us. He never fails us. So this came as a surprise. “*

*“Our faith is our protection. It acts as a shield against anything. It never shattered during troubled times*.*”*

All of these statements show the three means of supernatural protection that the participants have believed in. Karma (doing good things can result in good results later), God, and individual spiritual practice of praying have acted as the means of survival from COVID 19 infection. This was a strong perception that acted in their minds during the Pandemic situation.

There were participants who perceived the reason for acquiring COVID 19 to be supernatural to a certain extent.

*“I got this due to past sins in my previous lives. “*

*“God taught me to be aware of the sins. This is a way of showing me*.*”*

*“I have been following all the precautions as the doctors say. Our Karma can explain me succumbing to this disease*.*”*

These imply the viewpoint of supernatural causes, past sins and Gods’ punishment in contacting the disease. There was no ascertaining the disease into a biomedical model.

#### 2. Adherence to health care guidelines despite fatalistic beliefs

Although they had fatalistic ideas with regard to perceived COVID 19 risk, participants had adhered to health guidelines as much as they could. According to the participants, they had always worn face masks, maintained hand hygiene and maintained a 1-meter distance.

“Of course, I followed what doctors told me to do. With them and my good deeds, I thought I would be protected double from any ailment.”

“God helps the person who obeys their doctors. I wore masks and avoided crowded places. That way, God acknowledges me and protects me.”

The participants had not fully handed over the responsibility of their health to supernatural entities. They perceived that they had to care for themselves, and then the support from the supernatural will increase.

#### 3. Religious coping in COVID 19

The COVID 19 infected state seemed distorted the lives of infected persons fully. They had been treated in an isolated treatment care facility. Their families had been quarantined. There had been a varying level of stigma for COVID 19 infected people and their families. Such events lead to psychological turmoil. Religion had been found to be a tool in both practical and psychological coping with the distress associated with the Pandemic.

#### 4. Spiritual practices for coping with stress/ distress?

Spiritual practices seemed to have contributed to the study participants to cope with these situations. Following practices have been used by the study participants to reduce stress.

*“I am depressed. But, what to do? Lord Buddha has said that illness is inevitable. I think about it all the time*.*”*

*“I pray whenever I feel lonely. My family is alone. They have undergone a lot. I call them, and we pray to God together to give us strength to face what is in store. I know he will strengthen us*.*”*

*“When I feel sad, I will read the holy text. It seems like my hand was guided by God. Each time I touched the book, I was confronted with a consoling statement from the text*.*”*

All of the statement depicts high adherence to spiritual practices. They have been trying to cope up with the psychological effects of the Pandemic via these practices

#### 5. Fatalism as a positive force for self-care

Other than the real death situations, most of the study participants have entertained the fatalistic ideas of the inevitability of illness as a viewpoint to perceive life positively.

*“Well, ‘Atalodahama’ is said to govern the life of every being. If Lord Buddha got an illness, how can we be spared? This is the nature of ‘Sansara’. I meditate thinking about the*

*impermanence of life*.*”*

*“If you are alive, you have to face illnesses. Who can prevent that? This can happen to anyone*.*”*

*“God works in mysterious ways. This is one of the incidences He has taught me the value of praying. This is up to God to make me better. But I know he won’t fail me. When thinking about it I feel relieved*.*”*

Fatalism does not act as a force to feel hopeless in these scenarios. They have helped them to cope with the psychological distress through the mechanism of inevitability and a sense of supernatural protection.

## Discussion

In this context, the participants have perceived the risk of COVID 19 as low due to their spiritual protection and have attributed supernatural causes for contacting the disease.

Similarly, In sub-Saharan Africa, COVID 19 had been perceived mainly as a result of a supernatural phenomenon (war between Good and Evil) (16). Ali et al. and Malik have shown that COVID 19 is a test or punishment of God among Pakistan locals (17, 18). Similar points of view were observed among current study participants when ascertaining supernatural causes for Pandemic.

However, the participants of this study stated that even though even at the presence of supernatural support, there should be a commitment in public towards health guidelines adherence. In the present findings, religious faith acted as an augmenter for adherence to health guidelines. They have perceived that the faith had protection against COVID 19. But it has\d not hindered adherence to health guidelines. In contrast, the viewpoint based on faith has been found to be a negative factor for risk perception in COVID19, whereas in the African continent, mainly the man’s role in preventing the Pandemic is underestimated (16). Furthermore, there was a general rejection of *Public Health Measures* against COVID 19 in Africa(19). In Pakistan, the *Public Health Measures* such as lock-down was met with strong public resistance due to restrictions to participate in the mosque (20). The reason for these differences might be the religious, ideological differences between the two societies.

Especially Sri Lankan society is influenced by Buddhism, where the importance of ones’ own responsibility towards an effect is emphasized within the philosophy and practices (21). In contrast to that, Sub-Saharan African society is influenced by religions giving much importance to supernatural forces than free will with regard to health (22).

According to the present study participants the supernatural protection through religion was assumed worked against the Pandemic. Faith was considered a shield against COVID 19 among less than 50% of Catholics(23).While some form of divine protection was assumed to be obtained against COVID 19 through holy objects and spiritual practices in Mexico and Uganda(24, 25).

Wildman at el [Ma1] has stated that religion as a negative factor for pandemic outbreak control(26). However, our findings have emphasized the more positive impact of religion. Most of the study participants were Buddhists with Catholic and Muslim minorities. The basic cultural thinking patterns of the importance of an individual seem to have deeply rooted in religions practiced in this country, leading to more self-care behavior. This differs greatly from even the similar religions practiced elsewhere when taking this Pandemic as an example. The reason might be the concept of cultural religiosity, which explains the cultural affinity of the religiosity of an individual (27, 28).

Among the present study participants, the fatalistic notion that was associated with faith had acted as a positive reinforcement against distress and prompted to take public health guidelines as meritorious acts supported by a superior force. Thus, fatalism has affected positively in this context. In contrast, fatalism had been found to be negatively associated with compliance to public health measures when associated (29, 30) and not associated with religion. Fatalism during COVID 19 leads to anxiety in many examples (31, 32). The difference between these perceptions and the use of fatalism efficiently in this study can be related to the previously mentioned cultural importance of individual responsibility towards health or other aspects of life.

In this study, the study participants have used spiritual practices as tools to reduce distress and be positive minded. Many studies have reflected the importance of religious practices in different contexts in coping with stress associated with the Pandemic (7, 33, 34).

Furthermore, fatalism was used by this study participant as a positive force. However, fatalistic attitudes have been found to be having a negative impact on both psychological and physical health in many other types of research. It seems these have succeeded so far without succumbing the patients into a depressed state. This demonstrates the resilience provided by religion as a positive force.

Most perceptions described in this study shows the presence of semi-rational supportive views on COVID 19(35). They believe in the supernatural causation of the Pandemic yet adhere to the health guidelines. However, they never forgot their spiritual practices, entrusting their protective power. Ultimately those practices had auxiliary benefits. This reflects behaviors of the mid pathway not being extremely fatalistic yet adhering to opportunistic rituals.

## Data Availability

All data produced in the present study are available upon reasonable request to the authors

## Compliance with Ethical Standards

Ethical clearance for this project was obtained from the Ethical Review Committee of North Western Province Health Department

